# Quickly and simply detection for coronaviruses including SARS-CoV-2 on the mobile Real-Time PCR without treating RNA in advance

**DOI:** 10.1101/2020.08.06.20168294

**Authors:** Masaaki Muraoka, Yukiko Tanoi, Tetsutaro Tada, Mikio Mizukoshi, Osamu Kawaguchi

## Abstract

SARS-CoV-2 was reported to the WHO as an outbreak in Wuhan City, China on end of 2019, afterwards pandemic on the worldwide in 2020. The SARS-CoV-2 virus is less deadly, but far more transmissible. Therefore, it needs to detect and monitor quickly and simply on site to prevent SARS-CoV-2.

If detecting coronaviruses including SARS-CoV-2, the real-time RT-PCR method is sensitive and specific for the unique target, however, it must take long time and labour that RNA is treated in advance, transcribed and amplified. Therefore, referenced previously report, in this study, we modified various methods to prove hypotheses the followed.

Firstly, we hypothesized that real-time RT-PCR could be finished in very short time by the mobile real-time PCR device and one-step RT-PCR reagent. Secondly, we hypothesized that it was possible to perform RT-PCR utilizing the reagent as the above without RNA treatment in advance so called “direct”.

Firstly, it was able to detect the positive control RNA of SARS-CoV-2 for less than 13.5 minutes by primer-probe referring to the CDC. Moreover, each detection value varied in accordance with each concentration (This correlation coefficient R^2^ > 0.95). Secondary, it was possible to detect human coronavirus 229E with direct RT-PCR. Furthermore, each detection value varied in accordance with each titer (TCID_50_ / mL) of human coronavirus 229E (This correlation coefficient R^2^ > 0.95).

Considering the above, causing by utilizing the mobile real-time PCR device and the one-step real-time PCR reagent simultaneously following as: 1) It was possible to detect SARS-CoV-2 in very short time as compared to conventional method; 2) It was possible to detect human coronavirus quickly and simply with “direct”. For these reasons, we hypothesized that it is possible to detect SARS-CoV-2 quickly and simply by utilizing methods the above without treating RNA in advance. This hypothesis is our next try.

**STRENGTHS AND LIMITATIONS OF THIS STUDY:**

* This study developed it possible to detect the positive control RNA of SARS-CoV-2 more quickly than previously, however couldn’t try to detect the genetic RNA.
* This study proved clearly that the human coronavirus instead of SARS-CoV-2 could be detected simply without treating RNA in advance by the same method above.
* This study couldn’t try to utilize the human specimens because of our institution limited.
* This study could utilize the device and the reagents commercial and not especial.

## INTRODUCTION

Severe acute respiratory syndrome–coronavirus 2 (SARS-CoV-2) is a cause of coronavirus disease 2019 (COVID-19) outbreak, of which the World Health Organization (WHO) declared pandemic on March 11, 2020 [1]. COVID-19 was firstly reported to the WHO as an outbreak in Wuhan City, Hubei Province, China on December 31, 2019, afterwards has rapidly spreading [2]. SARS-CoV-2 included, human infectious coronaviruses have been known seven types. Four of the seven types (HCoV-229E, HCoV-HKU1, HCoV-OC43, and HCoV-NL63) are included among the known causes of common cold. Whereas, other three human coronaviruses (the Middle East respiratory syndrome coronavirus [MERS-CoV], SARS-CoV, SARS-CoV-2) are highly pathogenic and can cause severe diseases presented as acute respiratory distress syndrome (ARDS) characterized by dysfunctional immune responses and severe pulmonary injury [3, 4]. The SARS-CoV-2 virus is genetically closely related to SARS-CoV, but which is less deadly and far more transmissible than MERS-CoV or SARS-CoV [5]. Therefore, it has been needed with rapidity and on site to detect and monitor.

The conventional method of confirmation for SARS-CoV-2 is based on detection of sequences unique in the virus RNA by nucleic acid amplification tests (NAAT) such as real-time reverse-transcription polymerase chain reaction (RT-PCR). The viral genes targeted so far include the nucleocapsid (N), envelop (E), RNA-dependent RNA polymerase (RdRP), spike (S), and open reading frame 1 a/b (Orf1a/b), each of which primer and probe is various sequences [6, 7, 8, 9, 10, 11]. We recognize that the performance characteristics of PCR methods can vary with reagents, PCR programs, and devices. Therefore, if devices or reagents changed, with simpler and faster, it might be possible to detect.

Further, in quantitative RT-PCR, RNA extraction is required, followed by transcription, amplification and detection. Current RT-PCR typically require more than 60 minutes for RNA extraction, concentration and purification, and a further 90 minutes for transcription and amplification [12]. It is difficult to handle RNA not to be damaged and lost, possible to infect secondary and needed long time to detect. This might be due to the delay of the initial introduction of RT-PCR detection for SARS-CoV-2 in various places.

As already reported rapid detection of SARS-CoV-2 with using mobile real-time PCR device PCR1100 [10], we suggest “simpler and faster” method of RT-PCR in this paper as follows.

## MATERIALS AND METHODS

### Primer and Probe

Based on previously reported protocols of NIID [6, 7] and CDC [8, 9], and detection of HCoV-229E [13], PCR primers and probes for real-time RT-PCR detection were selected to target each N gene, and synthesised (Fasmac Co. Ltd., Japan) (Table 1).

**Table 1.**
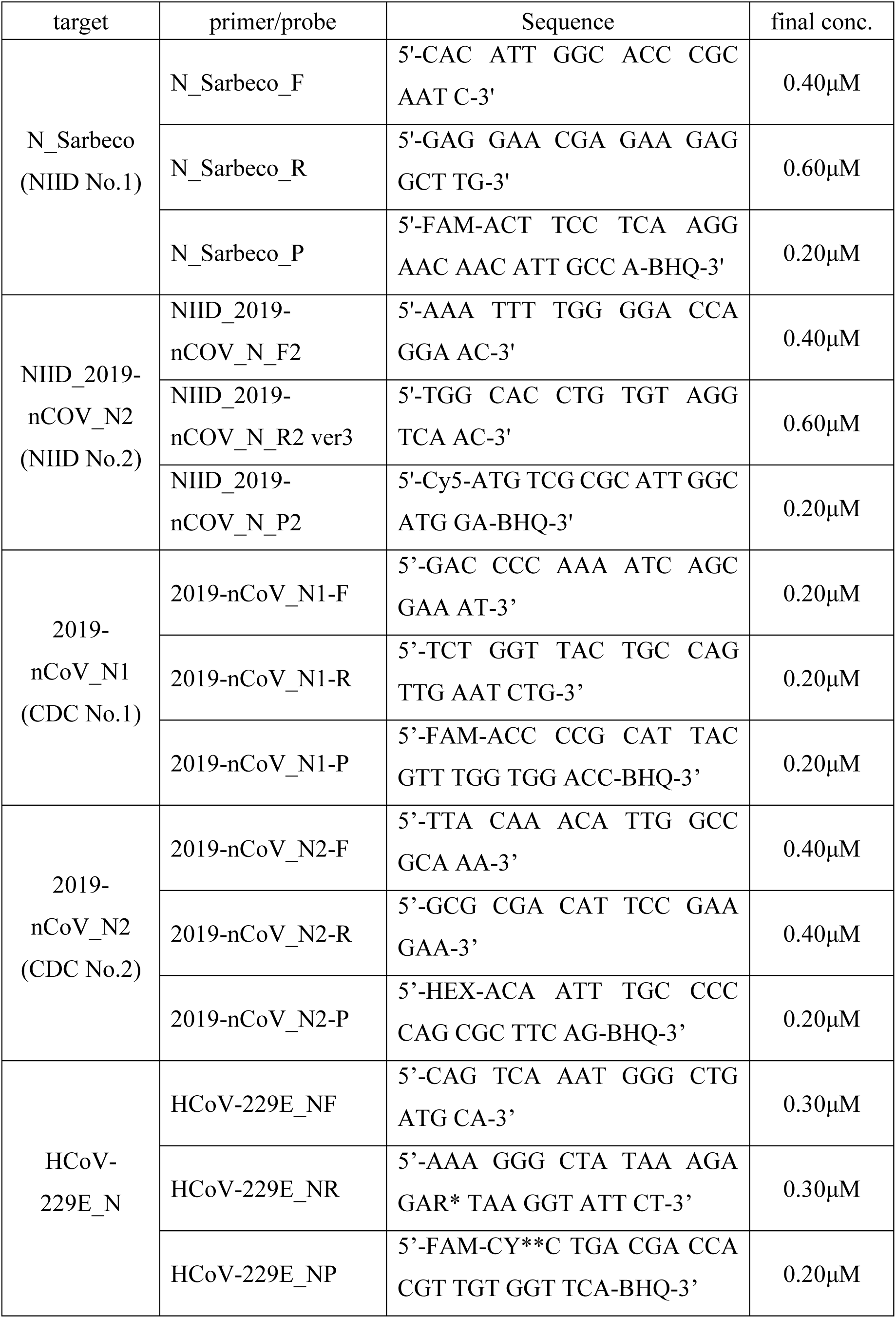
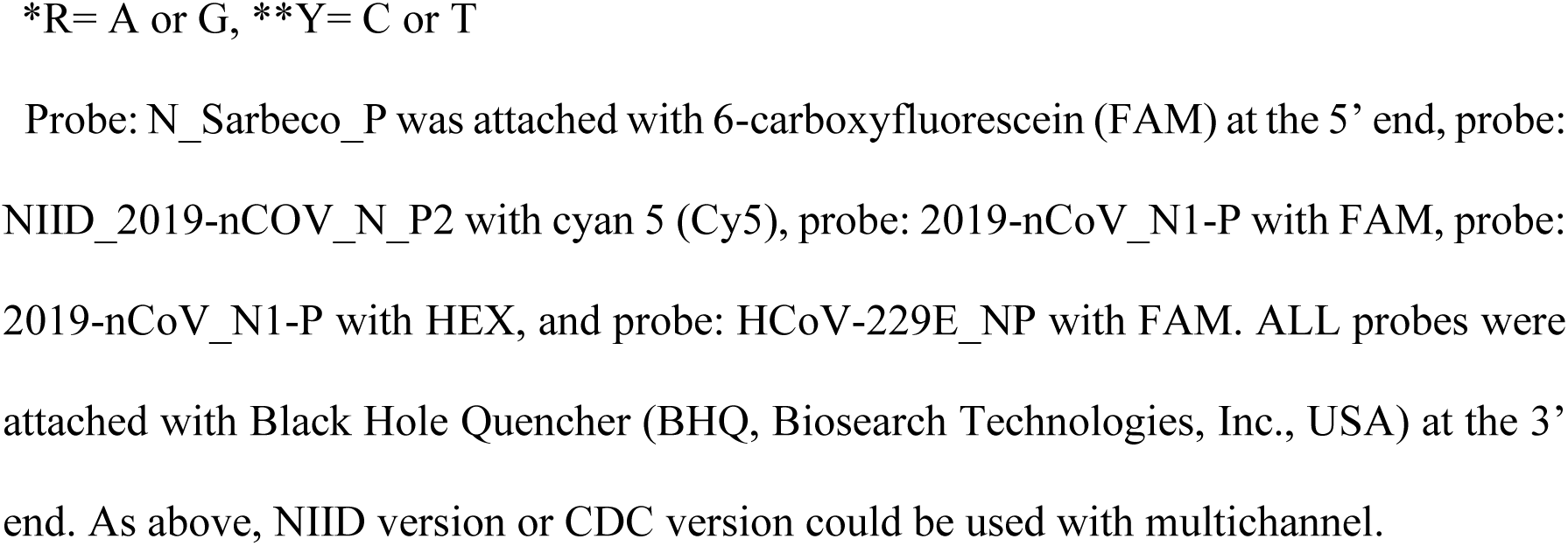
Selected primers and probes for the real-time RT-PCR of SARS-CoV-2 NIID and CDC, and HCoV–229E.

### Sample

Positive control RNA for SARS-CoV-2 against NIID version was kindly provided from NIID (for N primer probe Ver.2 and N2 primer probe Ver.2 / Ver.3), and against CDC version were synthesised (Fasmac Co. Ltd., Japan) based on each primer sequence and information of GenBank: MN997409.1 (Table 2).

**Table 2.**
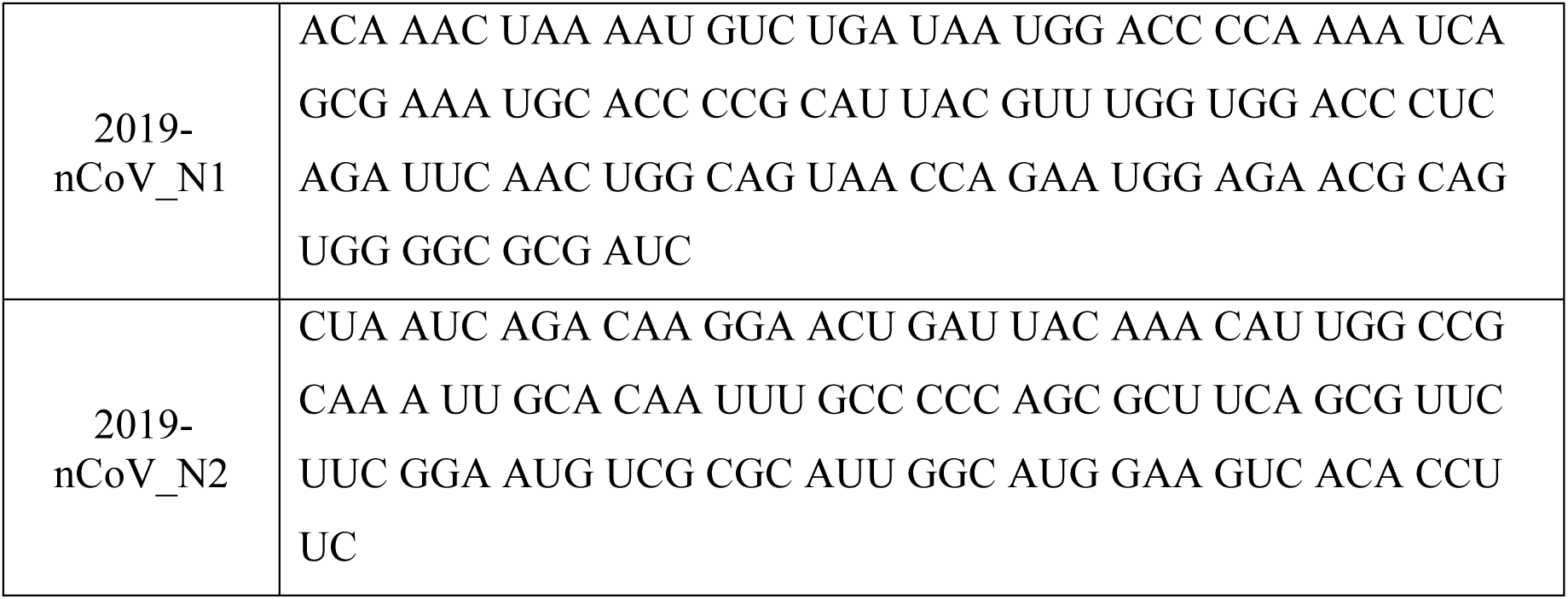
Sequences of positive RNA control for the real-time RT-PCR of SARS-CoV-2 of CDC version.

Each positive control RNA was applied 10-fold serial dilution with 10mM Tris-HCl (pH8.0 adjusted from 1M [Nippon Gene Co. Ltd., Japan] with water [UltraPure™ DNase / RNase-Free Distilled Water, Thermo Fisher Scientific Inc., USA]) included 10μg / mL carrier RNA (Ribonucleic acid from baker’s yeast, Merck KGaA, Germany). Further, each concentration of positive control RNA was assessed relatively with detection value of PCR1100.

The above, exposure to HCoV-229E is low-risk for healthy adults. The HCoV-229E virus is easily accessible for the biosafety level 2 (BSL2) laboratory. Therefore, HCoV-229E may be a good initial model for the evaluation against other coronaviruses, such as SARS-COV-2, the coronavirus that causes COVID-19 [14]. There we substituted HCoV-229E for SARS-2-CoV in direct RT-PCR. Human coronavirus 229E (HCoV-229E, VR-740™) was bought by certified non-profit organization biomedical science association in Japan (BMSA) from American Type Culture Collection (ATCC®, USA), afterwards, in cooperation of BMSA, it was able to be used. Because to proliferate HCoV-229E, it was infected with MRC-5 cells (CCL-171™, ATCC®), and cultured with EMEM medium (30-2003™, ATCC®) included 2% FBS (30-2020™, ATCC®) for 5∼8 days in 5% CO2 at 35 °C. When virus titer was grown up 8.9 x 10^6 Median tissue culture infectious dose (TCID_50_ / mL), only that supernatant without MRC-5 cells was stored at -80 °C before the assay. The supernatant included HCoV-229E was thawed in the assay, not treated in advance and used with crude as it was. To assess detection value of PCR1100 relative with titer of virus, the crude sample was applied 10-fold serial dilution with 10mM Tris-HCl (pH8.0, the above).

### Real-time RT-PCR

RT and PCR were carrying out in the one-step with using mobile real-time PCR device PCR1100 for all tests of this study. Briefly describe the composition of RT-PCR reagent, 1 μL of a sample (the crude or positive control RNA) was amplified in a 17 μL reaction solution containing 1x Reaction Buffer, 0.25 μL of RT Enzyme Mix, 1.0 μL of DNA Polymerase (THUNDERBIRD® Probe One-step qRT-PCR Kit, TOYOBO Co. Ltd., Japan), each final concentration of primer and probe for target detection (Table 2). With preliminary tested, each composition optimized (data not shown).

The RT-PCR conditions applied in this study were programmed as follows: RT incubation and enzyme activation were serially performed at 50 °C for 150 seconds, at 95 °C for 15 seconds respectively. Afterwards, for SARS-CoV-2 against NIID version: thermal cycling was then performed at 95 °C for 3.5 seconds (denaturation), and at 60 °C for 16 seconds (annealing and amplification) for fifty cycles. For SARS-CoV-2 against CDC version: thermal cycling was then performed at 95 °C for 3.5 seconds (denaturation), and at 60 °C for 7 seconds (annealing and amplification) for fifty cycles. For HCoV-229E: thermal cycling was then performed at 95 °C for 3.5 seconds (denaturation), and at 60 °C for 8 seconds (annealing and amplification) for fifty cycles. With preliminary tested, each condition was optimized (data not shown).

Incidentally, the procedure of PCR1100 was according to previously reported [15].

## RESULTS

### Detection of SARS-CoV-2 in positive control RNA

In NIID version, both No.1 and No.2 could be simultaneously detected with multichannel (Fig 1). As the result previously reported [7], No.2 was detected more sensitive than No.1, possible to determine 10 copies too. Each Ct value (detected on the PCR1100) was depended on the rate of dilution (No.1:R^2^ = 0.9982, No.2:R^2^ = 0.9969). When simultaneously detecting with two kinds of primer / probe sets, accuracy of results could be provided with higher reliability. It took only about 20 minutes per detection on the PCR1100 with using one step RT-PCR reagent (THUNDERBIRD® Probe One-step qRT-PCR Kit). So, it could be to detect 10 copies of positive control RNA only in 20 minutes.

**Fig 1.**
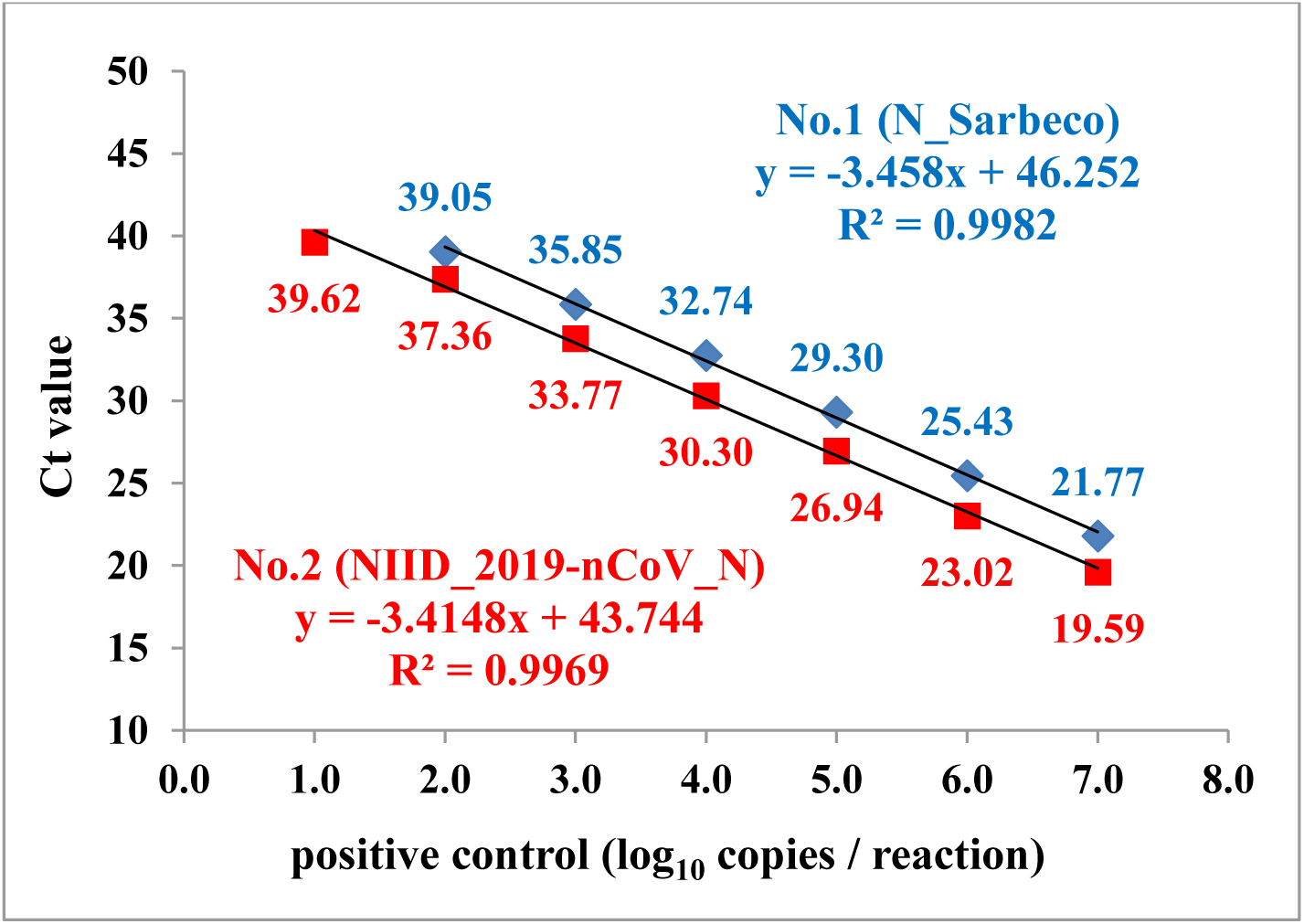
Correlation of copies number and Ct value when detection of positive control for NIID version on the PCR1100.

In CDC version, also, CDC No.1 and CDC No.2 could be detected with multichannel (Fig 2). Both No.1 and No.2 were detected in same sensibility, and possible to detect 10 copies too. Furthermore, each Ct value (detected on the PCR1100) was depended on the rate of dilution (No.1: R^2^ = 0.9996, No.2: R^2^ = 0.9983). As the same of NIID version, when simultaneously detecting with two kinds of primer / probe sets, accuracy of results could be provided with higher reliability. Furthermore, it took only about 13.5 minutes per detection on the PCR1100 with using the same one step RT-PCR reagent, that was even faster compared with NIID version. It could detect with at least10 copies of positive control RNA, besides it took only in 13.5 minutes.

**Fig 2.**
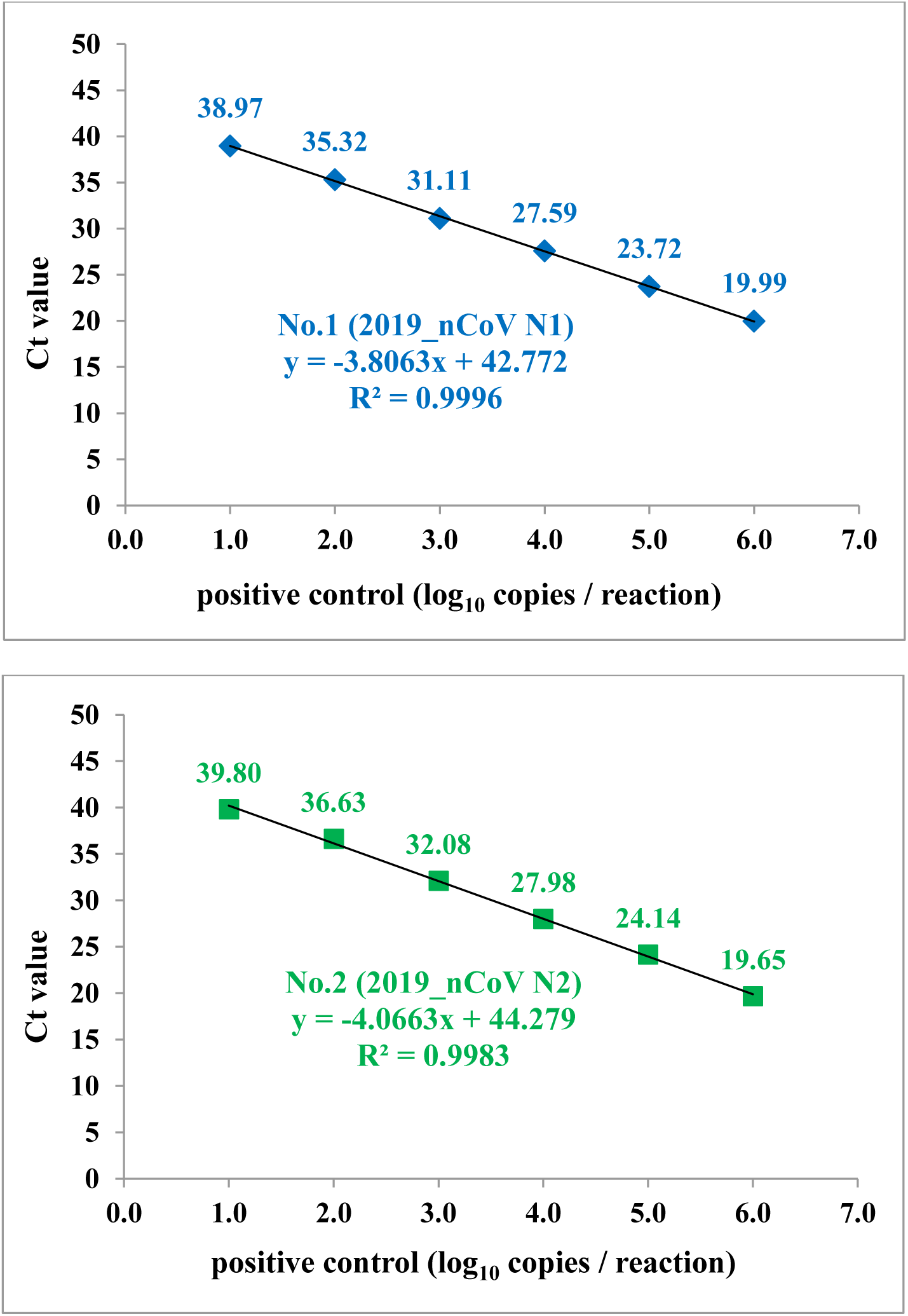
Correlation of copies number and Ct value when detection of positive control for CDC version on the PCR1100. The top is CDC version No.1 (2019-nCoV_N1), the bottom is No.2 (2019-nCoV_N2).

### Detection of HCoV-229E in crude state

Crude sample of HCoV-229E was applied 10-fold serial dilution, afterwards without treatment in advance and with one step RT-PCR reagent (THUNDERBIRD® Probe One-step qRT-PCR Kit) was tested in the RT-PCR on the mobile real-time PCR device PCR1100. As the result, without treatment for extraction, concentration and purification of RNA, it was possible to detect HCoV-229E at least 8.9 x 10^2^ TCID_50_ / mL (Fig 3). Furthermore, each Ct value (detected on the PCR1100) was depended on the rate of dilution, that is, virus titer (R^2^ = 0.9949). Ct value might be relatively indicated to virus titer.

**Fig 3.**
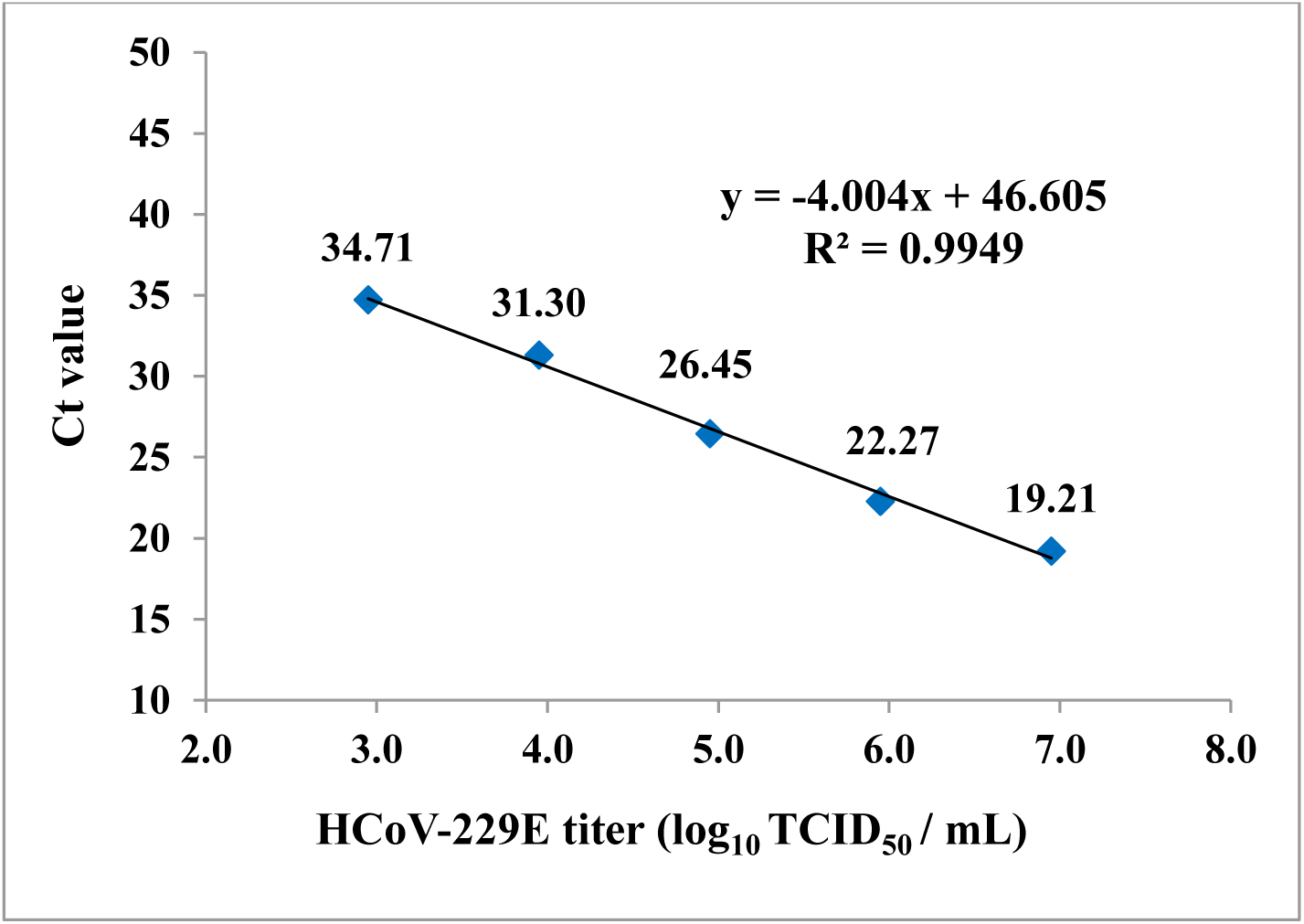
Correlation of virus titer and Ct value when detection of HCoV-229E on the PCR1100.

Also, it took only about 13 minutes per detection on the PCR1100 with using one step RT-PCR reagent the same above of SARS-CoV-2. On another real-time PCR device using normal two steps RT-PCR, it needed more than 90 minutes.

Therefore, it would be able to omit more than 135 minutes (60 minutes for RNA treatment, 75 minutes for RT-PCR) in total in detection of RNA virus compared with other current methods.

## DISCUSSION

Previously reported [10, 15, 16], with using the mobile real-time PCR device PCR1100, it could be to detect unique strain rapidly, and sensibility and specificity are equivalent to other devices. However, it would take 15 minutes at least to conclude RT-PCR for detection of unique RNA in any case. Furthermore, it was necessary for RNA in advance to adjust with extraction, concentration and purification.

First, this study addressed it to optimize each composition and PCR program according to previously reported [7, 10], a part of which modified to quickly carry out RT-PCR for detection of SARS-CoV-2 with using the mobile real-time PCR device PCR1100. With especially the unit of reverse transcriptase enzyme decreased and the time of reverse transcription shorted, sensibility of detection was improved (data not shown). Furthermore, as one step RT-PCR reagent, THUNDERBIRD Probe One-step qRT-PCR Kit (TOYOBO) was used, of which was possible and efficiently to carry out RT and probe’ real-time PCR simultaneously. This time, NIID and CDC version were selected for detection of SARS-CoV-2, both of which were unique for N-gene of SARS-CoV-2 and used in general [7, 8, 9]. As a result, in the NIID version No.2, CDC version No.1 and No.2, it was possible quantitatively to detect 10 copies or more of positive control RNA with using mobile real-time PCR device PCR1100. However, in the NIID, No.1 was able to detect only 100 copies or more, so No.2 was more sensitive than No.1 by 10 times. These results were about equal to previously reported [7, 10], that is, it might mean this method to be right adequate to detect positive control RNA for SARS-CoV-2. Furthermore, in both of CDC version by multichannel method, RT-PCR run could be finished in less than 14 minutes. That is, after RNA finished of extraction from each sample, this device would be quickly able to discriminate presence of target. This faster RT-PCR might be caused by combination of PCR1100 with RT-PCR reagent, of which was possible and efficiently to carry out RT and PCR simultaneously. However, regretfully, in our situation unable to deal with SARS-CoV-2, this method couldn’t be demonstrated possible or not for genome RNA as reported previously [7, 10]. Furthermore, primer / probe sets unique for gene of SARS-CoV-2 has been variously developed till now, so we would like to investigate about those sets if RT-PCR is able to be faster to more in the PCR1100.

Second, we tried it to modify total system to detect SARS-CoV-2 easier. When treatment in advance for RNA (mainly extraction, concentration and purification) it was required more than 60 minutes [12, 13, 17]. Therefore, in this study, we tried it to carry out RT-PCR without treatment of RNA in advance, so that, direct RT-PCR. We tested for direct RT-PCR, in that was used THUNDERBIRD® Probe One-step qRT-PCR Kit as one step RT-PCR reagent. With above, this reagent was possible and efficiently to carry out RT and probe’ real-time PCR simultaneously, furthermore as reason, described possible to NAAT from crude samples [18]. Additionally, because of our situation unable to deal with SARS-CoV-2, therefore we substituted HCoV-229E, low-risk for healthy, for SARS-2-CoV in direct RT-PCR this time. As a result, surprisingly, without treatment for extraction, concentration and purification of RNA, it was possible to detect HCoV-229E at least 8.9 x 10^2^ TCID_50_ / mL. Furthermore, Ct value could be relatively to virus titer. This time, Lower Limit of Detection (LoD) couldn’t show accurately because failed to measure less than 8.9 x 10^2^ TCID_50_ / mL. However, as approximate curve shown (y = -4.004x + 46.61), the LoD is supposed to less than 1 TCID_50_ / mL. Also, it took only about 13 minutes per detection on the PCR1100 with using one step RT-PCR reagent the same above of SARS-CoV-2. Therefore, it would be able to omit more than 135 minutes (60 minutes for RNA treatment, furthermore, 75 minutes for RT-PCR) in total in detection of SARS-CoV-2 compared with other current methods. Already some reported direct RT-PCR for SARS-CoV-2, it was necessary for especial reagent and additional treatment duration to direct RT-PCR [12, 19, 20, 21, 22]. So, we could suggest novel method in this paper, in which used PCR1100 device and commercial one step RT-PCR reagent only. However, our method must be assessed in real SARS-CoV-2, further validated using clinical specimens, which contained nasopharyngeal swabs and sputum. Furthermore, real-time RT-PCR reliable in saliva reported [23, 24], its specimen must be assessed too. On the other hand, if low virus in air could be collected, it might be able to detect quickly and easily on site by our novel method, and the result be following prevent from droplet infection.

Although the other four community-acquired HCoVs (229E, OC43, HKU1, NL63) are a common cause of common cold only, they are thought to cause pandemics and major outbreaks of probably more severe respiratory diseases when they initially crossed species’ barriers to infect humans decades and centuries ago [3, 25]. That is, it would be possible new coronaviruses to transfer again, therefore we are exposed to dangerous forever. On that time, the mobile real-time PCR could be it save time compared to earlier detection methods. Furthermore, we think that methods suggested in this paper should be suitable for various strains and it to be useful.

## STUDY DESIGN

When each sample was assessed in the real-time RT-PCR, a ten-fold dilution series of each was indicated with high relativity to Ct value on the mobile real-time PCR device PCR1100 (R^2^ = 0.99).

## Data Availability

The data that support the findings of this study are openly available at each URL each reference number in this manuscript.

## ACKNOWLEDGEMENTS

We thank Dr. Kazuya Shirato and Dr. Tsutomu Kageyama at the National Institute of Infections Disease in Japan (NIID) for positive control or technical comments, and Takashi Fukuzawa in an ex-employee at Nippon Sheet Glass Co. Ltd. for technical comments or experimental help.

## AUTHOR CONTRIBUTIONS

M. Muraoka conceived, designed experiments, performed experiments, analysed the data and wrote the manuscript. Y. Tanoi performed experiments and analysed the data. T. Tada wrote the manuscript. M. Mizukoshi conceived and provided specimens. O. Kawaguchi conceived and designed experiments. All authors also participated in the editorial process and approved the manuscript.

## ETHICAL STATEMENT

This study did not contain any studies involving human participants and animals performed by any of the authors.

## COMPETING INTERESTS

The authors declare that no competing interests exist. While M. Muraoka, T. Tanoi, T. Tada, and O. Kawaguchi were employed at Nippon Sheet Glass at the time of this study, in where all did not create a competing interest. Further, Nippon Sheet Glass had no involvement in this study.

## Notes

### Competing Interest Statement

The authors have declared no competing interest.

### Funding Statement

No external funding was received to me this time.

### Author Declarations

I did not have to be obtained necessary IRB and/or ethics committee approvals in this time.

### Summary of Updates

1) The erratum of name in the Author list was corrected. 2) The abstract, the introduction, and so on were corrected grammatical erratum.

